# Protocol for the PATHOME Study: A Cohort Study on Urban Societal Development and the Ecology of Enteric Disease Transmission among Infants, Domestic Animals, and the Environment

**DOI:** 10.1101/2023.05.26.23290617

**Authors:** Kelly K Baker, Sheillah Simiyu, Phylis J. Busienei, Fanta D Gutema, Bonphace Okoth, John Agira, Christine S Amondi, Abdhalah Ziraba, Alexis G Kapanka, Abisola Osinuga, Collins Ouma, Daniel K Sewell, Sabin Gaire, Innocent K Tumwebaze, Blessing Mberu

## Abstract

1.

**Introduction:** Global morbidity from enteric infections and diarrhea remains high in children in low- and middle-income countries (LMICs), despite significant investment over recent decades in health systems and global water and sanitation infrastructure. Other types of societal changes may be required to reduce disease burden. Ecological research on the influence of household and neighborhood societal development on pathogen transmission dynamics between humans, animals, and the environment could identify more effective strategies for preventing enteric infections.

**Methods and analysis:** The “enteric pathome” - i.e., the communities of viral, bacterial, and parasitic pathogens transmitted from human and animal feces through the environment is taxonomically complex in high burden settings. This integrated cohort-exposure assessment study leverages natural socio-economic spectrums of development to study how pathome complexity is influenced by household and neighborhood development. We are enrolling under 12-month-old children in low- and middle-income neighborhoods of two Kenyan cities (Nairobi and Kisumu) into a “short-cohort” study involving repeat testing of child feces for enteric pathogens. A mid-study exposure assessment documenting infrastructural, behavioral, spatial, climate, environmental, and zoonotic factors characterizes pathogen exposure pathways in household and neighborhood settings. These data will be used to inform and validate statistical and agent-based models that identify individual or combined intervention strategies for reducing multi-pathogen transmission between humans, animals, and environment in urban Kenya.

**Ethics and dissemination:** The protocols for human subjects’ research were approved by Institutional Review Boards at the University of Iowa (ID - 202004606) and AMREF Health Africa (ID - ESRC P887/2020), and a national permit was obtained from the Kenya National Commission for Science Technology and Innovation (NACOSTI) (ID# P/21/8441). The study was registered on Clinicaltrials.gov (Identifier: NCT05322655). Protocols for research on animals were approved by the University of Iowa Animal Care and Use Committee (ID 0042302).

**STRENGTHS AND LIMITATIONS OF THE STUDY:**

- This cohort-exposure assessment study will provide new evidence on the nature of household and neighborhood developmental strategies that are most effective at preventing critical multi-pathogen transmission pathways among humans, animals, and environment in cities of low- and middle-income countries.
- We study middle class households and neighborhoods to test counterfactual theories about meeting basic developmental standards to reduce pathogen transmission.
- Our data collection uses objective methods to comprehensively document socioeconomic, weather, infrastructural, spatial, behavioral, environmental, zoonotic, and human data, including use of both selective culture and molecular methods to characterize pathogen community patterns.
- The observational study design is vulnerable to unmeasured confounders.
- The living conditions in middle-class households and neighborhoods may not offset hygiene conditions in the overall urban environment enough to alter enteric pathogen transmission patterns.

## Introduction

### Study Rationale

In the past half century, advances in access to health care and early childhood vaccines and therapies both in high income countries (HICs) and in low-to-middle income countries (LMICs) have successfully reduced global mortality from diarrheal diseases. According to Global Burden of Disease (GBD) data, diarrhea mortality in children under five years decreased by 59·3% (from 173·3 per 100⍰1000 to 70·6 per 100⍰1000) between 2000 and 2016, largely in South Asia and sub-Saharan Africa where fatality rates were highest [1]. However, reductions in diarrhea incidence have lagged with only a 12·7% reduction (from 2·0 per child-year to 1·75 per child-year) over the same 16 years, and large differences remain between incidence in children in HICs (0.58 per child year) versus regions like Latin America and sub-Saharan Africa (2.82 cases and 2.37 cases per child-year, respectively). Typhoid and paratyphoid enteric fever cases in endemic areas also remain high at 14.3 million cases per year [2]. Asymptomatic infection prevalence is even higher than symptomatic rates [3], and symptomatic and asymptomatic infections can elevate a child’s susceptibility to co-infection [4], enteric dysfunction and malnutrition [5, 6], and long-term cognitive and developmental stunting [7, 8]. Greater investment in disease prevention and control programs that protect children from pathogen exposure will be needed to accelerate the reduction in global enteric disease burden and achieve health equity between HIC and LMIC populations.

At the end of the 19^th^ century, typhoid fever, cholera, and diarrhea epidemics routinely plagued cities like London and Philadelphia in today’s HICs [9]. Annualized death rates were quite similar to rates in LMICs today, although with stronger seasonal patterns [10]. Death rates dramatically decreased in the late 19^th^ to early 20^th^ centuries, even as urban populations were growing rapidly and well before the development of vaccines [11]. Development of municipal piped water and wastewater systems contributed to these health transitions, although inference from these studies is limited by the lack of model adjustment for confounding from other causal pathways (food safety, handwashing, housing quality) [12-14]. Water, Sanitation and Hygiene (WASH) on has occupied a central strategy in global development policy over the last fifty years for reducing global diarrhea burden and resulted in impressive increases between 1990 and 2015 in global population with access to household drinking water and to a lesser extent sanitation infrastructure. These investments have improved quality of life globally, but as stated above have not generated expected declines in diarrheal incidence in LMICs [15]. Randomized controlled studies reporting relatively small benefits from household-level improvements in WASH and recent demographic studies raise questions as to what other conditions are required for disease control [16-20]. Achieving population-wide water and sanitation coverage, infrastructure maintenance, and chlorination of a water supply may be necessary, but not sufficient for disease control [19-24].

Control of enteric pathogen transmission may require more wide sweeping overhauls of societal sanitary conditions beyond drinking water and sanitation systems. The building of water and sanitation systems in HIC cities was just one piece of a much larger societal reformation that included parallel improvements in health systems, housing quality, waste disposal laws, awareness of personal hygiene, growth of a middle class, and better nutrition [25]. The scientists who were directly observing health transitions between 1850 and 1950 proposed diverse theories for the collapse of enteric disease epidemics. Some studies noted improvements in water sources in Glasgow and London correlated poorly with observed declines in diarrhea rates over time, with major decreases in disease occurring decades after improved water sources and decades before sewerage [10, 26]. Contamination of milk sources was considered a cause of typhoid and scarlet fever outbreaks [27, 28]. Widespread regulation of milk pasteurization also emerged in the late 19^th^ century and has been attributed with decreases in diarrhea case rates in Philadelphia and New York City in the 1900’s [29, 30]. Others suggested sharing of unsanitary living conditions rather than sewerage problems caused typhoid fever cases [31]. One comprehensive study examined housing quality, shared living conditions, food supply and type of water and toilet in early 1900’s Mansfield, England, which had a seasonal diarrhea prevalence of 10% despite high sewered sanitation and water supply coverage [32]. It supported the premise that shared housing and unhygienic household habits contributed most to diarrhea prevalence and noted that middle class households in otherwise crowded areas with poor neighborhood hygiene were not protected from infection.

Evolutionary biology evidence suggests that the establishment of permanent agricultural population centers around 10,000 years ago triggered pre-industrial epidemiological transitions that *increased* pathogen adaptation to humans and multi-parasitism in human populations. Migration and urbanization further concentrated animals, humans, and their waste into relatively small areas, increased pathogen host adaptation, and increased rates of within-species and across-species transmission of many pathogenic species [33]. Over time the number of households in HICs choosing to own animals and the types of animals own has shifted as a byproduct of changing lifestyles, technology, and limited space and need for animal rearing. The importance of animals to urban industry has also shifted. Brownlee and Young (1922) reported strong consensus at scientific society meetings with the theory that the replacement of horses and gravel road surfaces with motorized vehicles and tarmac prior to 1910, reduced visible animal feces in the streets and coincided strongly in time with disease declines. Strong temporal correlation between decreases in annual issued licenses for horse-drawn vehicles in English and Welsh cities and declines in diarrheal deaths provides support for that premise [34]. Fly control may have further enhanced the community hygiene-driven declines in typhoid fever and diarrheal deaths [35]. Changes in contact between humans, work animals, and other domestic species that prevented zoonotic transmission could also explain early epidemiological transitions.

Elimination of cholera, typhoid fever, and other enteric disease may have required a combination of societal developmental changes that collectively attained a standard of environmental hygiene capable of preventing transmission of infectious agents– essentially a microbial community effect. Unfortunately, evidence on which aspects of development had the greatest impact on enteric disease ecology during historical transitions cannot be disentangled because records on the timing, rate, and scale of coverage of most societal development activities, and personal adoption of hygiene behaviors are fragmented and poorly documented [36]. Which persons benefitted from those improved living conditions is also poorly recorded, requiring studies to rely upon aggregated health records until the middle of the 20^th^ century [14]. Records on outcomes are predominantly death records for cholera and typhoid due to inconsistent health care seeking and recording of non-fatal cases and investigations of other enteric causes of diarrheal deaths. Contemporary studies examining societal development and multi-decade enteric disease rates in countries that have recently undergone epidemiological transitions would be more culturally relevant, but such studies are also lacking for many of the same data availability challenges.

### Objectives

A study combining Planetary Health and One Health approaches to examining ongoing health transitions in high burden countries of the 21^st^ century could better inform which developmental factors most influence enteric disease prevention and control for prioritizing health program policy. To address this knowledge gap, we describe the scientific rationale, hypotheses, and design for the <u>Pa</u>thogen <u>T</u>ransmission and <u>H</u>ealth <u>O</u>utcome <u>M</u>odels of <u>E</u>nteric Disease (PATHOME) study supported by the National Institutes of Health (NIH) through the Evolution and Ecology of Infectious Disease funding mechanism (1 R01 TW011795, years 2020-2025). The objectives of PATHOME are to (1) characterize interactions among enteric pathogenic agents in infants, domestic animals, and the environment and the dynamic interactions between infants, caregivers, animals, and environmental materials across space and seasons; and (2) develop statistical and computational methods to examine this complex disease system and predict which social and environmental developmental improvements best prevent multi-pathogen transmission in urbanizing areas of a high disease burden country. Our overall hypothesis is that joint modeling of “enteric pathome” agents (i.e. the microbial communities of viral, bacterial, and protozoan pathogens transmitted by human and animal feces) across households and neighborhoods that represent contrasts in urban societal development will show that development leads to lower pathogen-specific detection frequencies, and thus evolution of the pathome from complex to simple microbial community structure distributions in humans, animals, and the environment (Figure 1). Our study design is motivated by several fundamental science theories (Table 1).

**Table 1.** Study Design Strategies and Scientific Rationale for the PATHOME Study.

| Study Design Strategy | Rationale |
| --- | --- |
| Recruit from neighborhoods representing low to moderate levels of societal development to test counterfactual hypotheses that high thresholds of basic development can reduce pathogen transmission | The rapid growth in middle-class urban households in LMICs globally can be leveraged to compare enteric pathogen exposure and health patterns across a socioeconomic developmental spectrum to identify the conditions that most influence enteric disease transmission. While increases in household socioeconomic status are known to decrease the risk of pediatric diarrhea in LMICs, most studies have been based in low-income populations. Evidence from middle class neighborhoods testing is critical for identifying tipping points in transmission and accurately predicting potential program impact on health outcomes. |
| Apply a One Health approach that includes zoonotic and environmental health to identify all sources of pathogen transmission | Domestic and sylvatic animals are susceptible to infection from many of the same enteric pathogens and may have more widespread carriage than humans due to less frequent disease surveillance and treatment. Wildlife are considered major drivers of novel emerging zoonotic disease outbreaks, but human interactions with domestic species may be more important in sustaining endemic zoonotic burden because the higher frequency of human-animal interaction increases human contact with zoonotic vectors. Keeping animals for food, pest control, and companionship are common practices in most urban and peri-urban LMICs settings and these animals are also often allowed to freely roam and forage for food in household and neighborhood areas which can spread feces with pathogens across the environment [37-39]. Improvements in sanitation coverage does not reduce animal feces pollution in the environment so data collection on safe feces management must account for infection prevalence in animals, animal hygiene practices, and human-animal contact. A similar study to this one in Nairobi detected human-animal *E. coli* genotype sharing but found sharing between humans to be more common [40]. Pathogen sharing is also influenced by exposure dose, type and effectiveness of microbial virulence genes, |
|  | and host anti-pathogen immunity, so within- and between-host pathogen sharing patterns may not be the same as for non-infectious *E. coli*. |
| Rely upon a microbial ecology approach to detect pathogen diversity at the human-environment-animal interface to identify pathogen sources and transmission dynamics | Studies in urban Kenya and elsewhere have demonstrated taxonomic diversity in enteric pathogens in humans [3, 18, 41], animals, and environment [39, 42, 43] in areas where open defecation, dumping of untreated sewage, and unsanitary animal management practices are common [44]. Taxonomic diversity varies by exposure pathway, and multi-pathogen exposure risks may be elevated in infants living in underdeveloped houses with dirt floors, shared latrines, and domestic animals. Evidence on pathogen diversity in middle-class neighborhoods is unknown, although theoretically should be low if moderate to high thresholds of neighborhood infrastructure and hygiene have been met. Identifying intervention strategies that best reduce multi-pathogen transmission could enable advocacy for improved investment in high impact developmental and public health programs. |
| Use a community-based enrollment design and objective measures of pathogen presence or absence to detect transmission dynamics that are unconfounded by host type, health status, and health care utilization behaviors | Reliance upon diarrhea symptoms as an indicator of exposure can lead to biased study designs that presume health care seeking cases are exposed while asymptomatic controls are presumed unexposed. The relationships between ingestion of a pathogen, gut infection, and progression to symptomatic status are influenced by many factors, such as type of pathogen, infectious dose, host immunity, comorbidities, and breastfeeding. Asymptomatic carriage of enteric pathogens is common in endemic areas [3], and these individuals can still pose a transmission risk to others, but typically go undetected due to lack of symptoms. Evidence from cities in the United States in the 1930s and 1940 suggests that city development decreased both symptomatic and asymptomatic *Shigella* infections [9]. Combining community-based sampling with objective measures of pathogen presence in infants, animals, and environmental samples avoids misclassification of exposed asymptomatic subjects as unexposed. |
| Enroll children between 0 to 12 months of age to improve detection of transmission dynamics | A high pre-existing pathogen detection prevalence makes it more challenging to distinguish between persistent or pre-existing infections versus new infections and increases the chances that pathogen detection in children is confounded by pre-existing host immunity or disease conditions. Large studies of enteric pathogen detection in under- five children consistently show that detection and co-detection of enteric pathogens in endemic areas climbs rapidly within the first months of life [45, 46]. Study of children between birth and twelve months leverages the dynamic increases in prevalence rate in this age group for identifying early causes of population disease transmission. |
| Use a short-cohort, repeat measures design that matches a natural timeline of exposure and infection to improve detection of transmission dynamics | For most enteric pathogens, the timeline between known exposure and confirmed infection is between 1 to 5 days. The timeline between infection and recovery varies from a few days to a few weeks. Pathogen carriage and diarrhea lasting more than 14 days is considered persistent and unusual. A cohort study that spaces human health assessment by 30 days or more cannot detect exposure-outcome relationships or confirm that concurrent pathogen detections over a month or more are persistent carriage versus new exposure events. Given known temporal pathogen-host disease trends, the dynamic rate of infections in infants in Kenya, and the evidence of widespread pathogen contamination in low-income urban neighborhoods, a 14-day short cohort design with repeat diagnostic testing for enteric pathogens in infants could detect |
|  | transmission events and monitor their progression. Assessing pathogen presence in infants at enrollment can improve screening for post-enrollment pathogen exposures. |
| Apply a Planetary Health approach to characterize household and community socio-economic and infrastructure development on enteric pathogen transmission | Natural societal development involves improvements in many living conditions that are inherently correlated, like water and sanitation access, flooring, ability to buy safer foods and soap for handwashing, and possibly changes in zoonotic exposures. The rate with which households and neighborhood improve their circumstances can vary creating spatial variability in when and where hygiene conditions are achieved. If human and animal feces containing enteric pathogens are discarded in drains or lots in the neighborhood, it can be carried by human and animal foot traffic or flood water into the home impacting household hygiene conditions and exposure of children [43, 46, 47]. Children also can be exposed to neighborhood soil, water, objects, and animal feces containing enteric pathogens during play or movement in residential environments outside the household [42, 43]. Understanding how changes in socio-economic and built environment conditions influence population health dynamics in the present could improve the future sustainability and health effectiveness of urban planning policies [48]. In the context of enteric disease control, evidence is needed on how household and community development and the spatial exposures of infants influences the collapse of enteric pathogen transmission. |
| Select enteric pathogen indicators that are likely to occur dynamically over space and time in the study population for discriminatory power | Pathogens selected for characterization of pathome transmission were chosen based upon evidence that (1) population prevalence of the pathogen in diarrheal and asymptomatic infants changes between 0 and 12 months of age in Kenya [3]; (2) pathogen detection in humans, animals, and/or environment is a sensitive indicator of unhygienic conditions; (3) domestic animals and environmental reservoirs (e.g., soil and water) contribute to pathogen transmission; (4) methods exist or can be adapted to isolate organisms and validate pathogen presence in human, animal, and environmental materials. |

**Figure 1.**
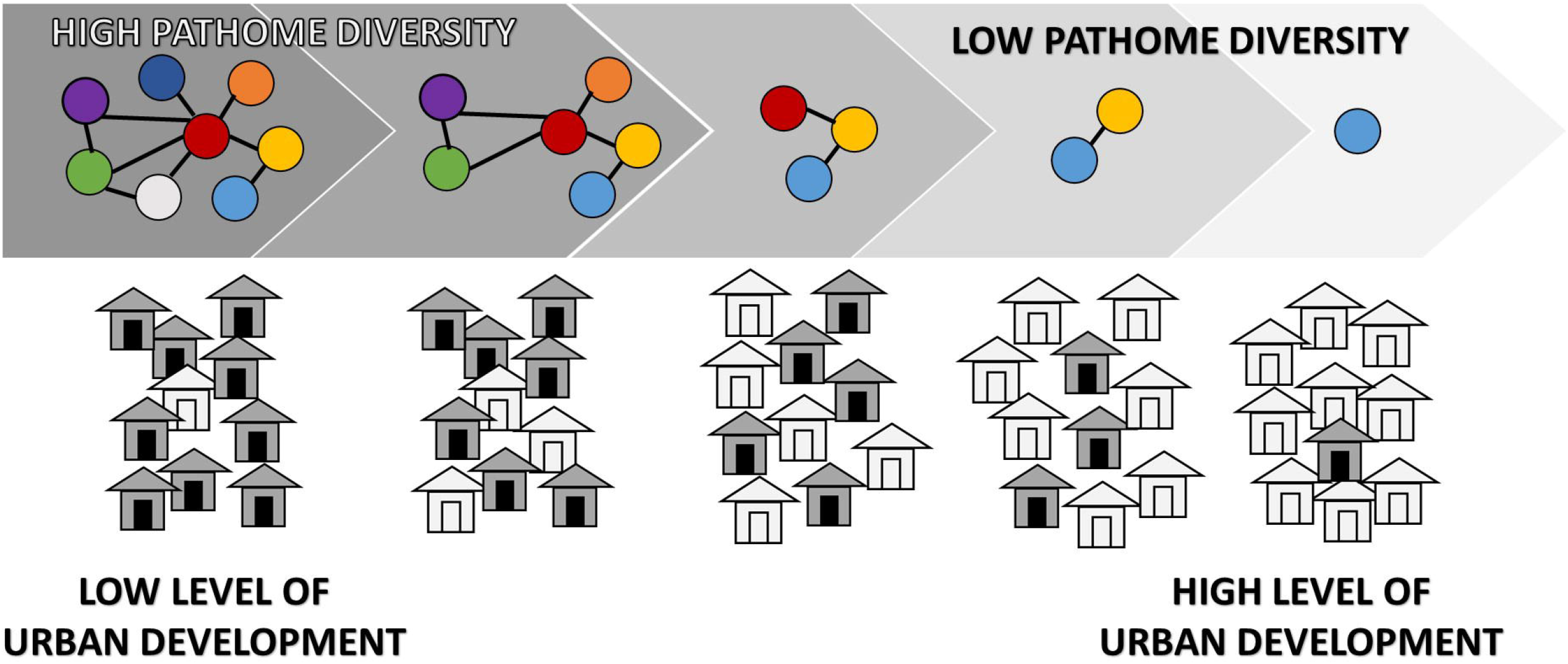
Conceptual Model of the Enteric PATHOME Study. Societal development of household water, latrines, flooring, animal health, and hygiene conditions and neighborhood infrastructure (drainage) and hygiene (animal penning, waste regulations) conditions triggers an evolutionary change in the enteric PATHOME from taxonomically complex to simple microbial community detection patterns in humans, animals, and the environment.

## METHODS AND ANALYSIS

### 1. Study setting

The study is situated in low-income and middle-income communities in two different Kenyan cities, Nairobi (population 4.4 million persons) and Kisumu (population ∼610,000 in 2019) where our team has performed prior health research [43, 45, 49-60]. The prevalence of diarrhea in children in Kenya is high with most cases occurring in children under 3 years of age, and roughly 25% of children are severely stunted [61]. However, diarrhea prevalence is much lower in Kenya’s rapidly growing middle class [59], indicating that epidemiological transitions in enteric disease risk are occurring along low-to middle-income neighborhood gradients. Communities in each city were selected based upon similarity in population’s ethnic and cultural characteristics but with distinct differences in overall community developmental conditions. Environmental hygiene indicators also differ by neighborhood socioeconomic status and thus are expected to be different in the two target cities. Private toilets are the norm in middle-income neighborhoods, while the majority of households in low-income areas rely upon shared latrines for disposal of human feces and a quarter of households dispose of feces in open defecation sites, drains, or waste dumps [62]. Low-income households [46, 62] and neighborhoods [39] are heavily polluted with indicator bacteria and enteric pathogens, while contamination of middle-income neighborhoods is unknown. Half of households in Kisumu keep domestic animals [37], while animal ownership is less common in low- and middle-income Nairobi neighborhoods. While development or the lack thereof may be uniform across communities, in urban settings there may be outlier households that differ from their neighbors, such as relatively wealthier households in low-income communities or relatively poorer households in middle-income communities. The communities selected for this study offer the opportunity to enroll these types of households and study how a spectrum of household conditions might influence transmission dynamics in the context of broader community conditions.

### 2. Study Design

To capture exposure-outcome relationships reflecting pathogen transmission dynamics, we are conducting a prospective 14-day cohort epidemiological study with repeat health outcome assessment and integrated exposure assessment data collection. We are recruiting 32 children in 4 age groups (<3 months, 3 to <6 months, 6 to <9 months, 9 to 12 months) on a rolling basis over 2 consecutive years across wet and dry seasons (12 months) in low-income and middle-income neighborhoods of Kisumu and Nairobi for a total of 248 households with complete data assuming a 10% attrition rate (Figure 2). Enrollment in Nairobi spans November 2021 to November 2022, with the last participant follow-up in December 2022. Enrollment in Kisumu spans late March 2023 to February 2024 with the last participant follow-up expected in March 2024. Participant recruitment follows a standardized schedule of sixteen households over a four-week period, eight each from the low- and middle-income neighborhood, followed by a one week break in enrollment for neighborhood-level observations and environmental sampling. This five-week cycle is repeated 12 times or until sample size requirements for each neighborhood are met.

**Figure 2.**
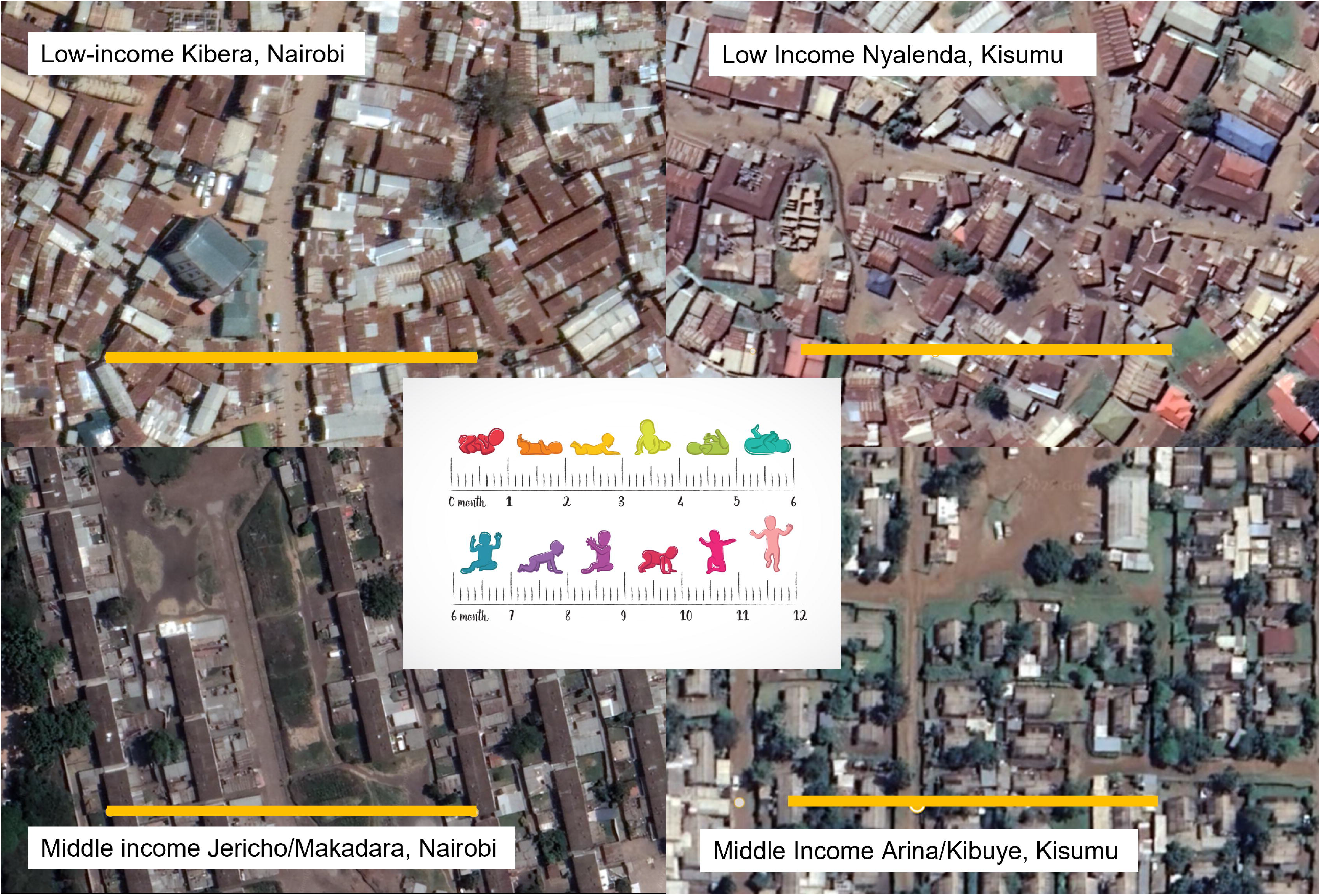
Low- and middle-income study neighborhoods in Nairobi and Kisumu, Kenya, where households with 0-to-12-month of age children are enrolled into the PATHOME study. Yellow bars represent 100 meters distance. Image licensed through Vectorstock Standard License.

### 3. Eligibility, Participant Recruitment, and Enrollment

Eligibility is defined as infants between 0 and 12 months of age, as verified by birth registration card. Infants with disabilities that would impact normal behavior are excluded. Study of infant age groups also provides an opportunity to examine pathogen transmission dynamics related to development of motor skills that allow infants to explore their environment [61]. Participants are identified from study neighborhoods by extracting infant age and the primary caregiver’s name and contact information from monthly surveillance records collected routinely by Kenyan Ministry of Health (MOH) Community Health Volunteers (CHVs) in each neighborhood. Per their standard practice, CHVs approach all households within their area of work with pregnant women or young children to ensure they receive health information and have support to access health care. Recruitment data is compiled as separate lists for the low- and middle-income neighborhoods of each city. When CHVs identify potentially eligible households, they notify study staff of the African Population Health and Research Center (APHRC) who follow up with households in the company of the CHV to verify information about age-eligible children.

The information sheet/form explains the study activities and the purpose for them, as well as risk and benefits, costs for participation (none), right to withdraw, and compensation. The adult/primary caregiver is informed about all study activities by the field staff who read the information on the form to the caregiver in English or Swahili, in the presence of their CHV as a witness, and answers any questions they may have. Consent forms are signed by the caregiver with signature or thumb print and by their CHV witness if the caregiver is not in a position to sign themselves. A copy of the information sheet and consent form is provided for the caregiver’s permanent records. After informed consent is completed, the study team schedules a time to begin data collection and provides the caregiver with a package of size-appropriate diapers with instructions to collect infant feces specimens over the subsequent two weeks. Collection of a baseline or Day 1 feces sample is required for continued enrollment in the study. Should consent or an enrollment feces sample not be provided, the research team and CHV identifies another infant of same age group and neighborhood from the list for recruitment. Biological sex of the infant is not an enrollment criterion and does not influence recruitment although we expect the sex of the study population to be representative of natural sex distributions in urban Kenya. Households are provided one pack of diapers and flour and sugar to make porridge (items identified by caregivers in a pilot study as being highly desired that directly benefit the child) as compensation for their time at the end of enrollment (approximately 6 USD).

### 4. Study Outcomes

The <u>primary outcomes</u> of the epidemiological study are detection and co-detection of specific types of enteric pathogens in infant feces during the enrollment period. The three approaches for examining pathogen detection are:

1. Pathogen prevalence - the proportion of infants with a specific type of enteric virus, bacteria, or parasite detected in their feces at enrollment (Day 1).
2. Pathogen incidence –the proportion of infants who have a specific type of enteric virus, bacteria, or parasite detected in their feces in the 13 days after enrollment that was not present in their feces at enrollment (Day 2 to 14).
3. Pathogen diversity – the number of unique types of enteric virus, bacteria, or parasite detected in infant feces at enrollment (Day 1).

The study is evaluating the following <u>secondary outcomes</u>:

1. 14-day prevalence of prospective self-reported diarrheal symptoms - proportion of caregivers who report an enrolled infant had 3 or more loose feces, with or without blood or mucus, in any 24-hour period during the enrollment period (Day 1 to 14).
2. 7-day prevalence of self-reported diarrheal and other symptoms prior to enrollment - proportion of caregivers reporting at enrollment that in the past 7 days the enrolled infant had symptoms of: malaise, vomiting, difficulty in breathing, unusual lack of appetite or willingness to take liquids, fever, runny nose, cough, rash (pre-Day 1).
3. Preterm birth prevalence - proportion of infants born before 37 weeks gestational age (Day 1).
4. Stunting prevalence - proportion of infants whose length-for-age z-score (centimeters) is −2 or more standard deviations away from the international standard during the enrollment period (Day 3 and 7).
5. Wasting prevalence - proportion of infants whose weight-for-age z-score (kilograms) is - 2 or more standard deviations away from the international standard (Day 3 and 7).
6. Mid-upper arm circumference (Day 3 and 7).

The study is evaluating the following Intermediate Pathogen Exposure Outcomes:

1. Frequency of child exposure behaviors - number of infant contacts with caregivers, other children, domestic animals, and the environment per unit of time (Day 2, 3)
2. Presence and concentration of *E. coli* and pathogenic sub-types, *Salmonella enterica, Shigella spp*., *Campylobacter jejuni/coli*, and *Listeria monocytogenes* in household (Day 2) and public (monthly) environments.
3. Prevalence and concentration of enteric viruses, bacteria, and protozoan pathogens in feces of domestic animals (Day 2).

### 5. Sample Size Determination

Sample size calculations aimed to ensure enough observations to reliably estimate the model parameters and limit the set of plausible parameter values to a reasonably narrow range. Towards this, we considered the set of models to be used to examine the relationship between exposures and multi-pathogen infections and ensured that we achieved 11 to 21 observations per parameter, depending on the model.

### 6. Data Collection

PATHOME methods are summarized below and in Figure 3, with detailed protocols provided sequentially in Supplemental Materials and at https://pathomelab.github.io/.

**Figure 3.**
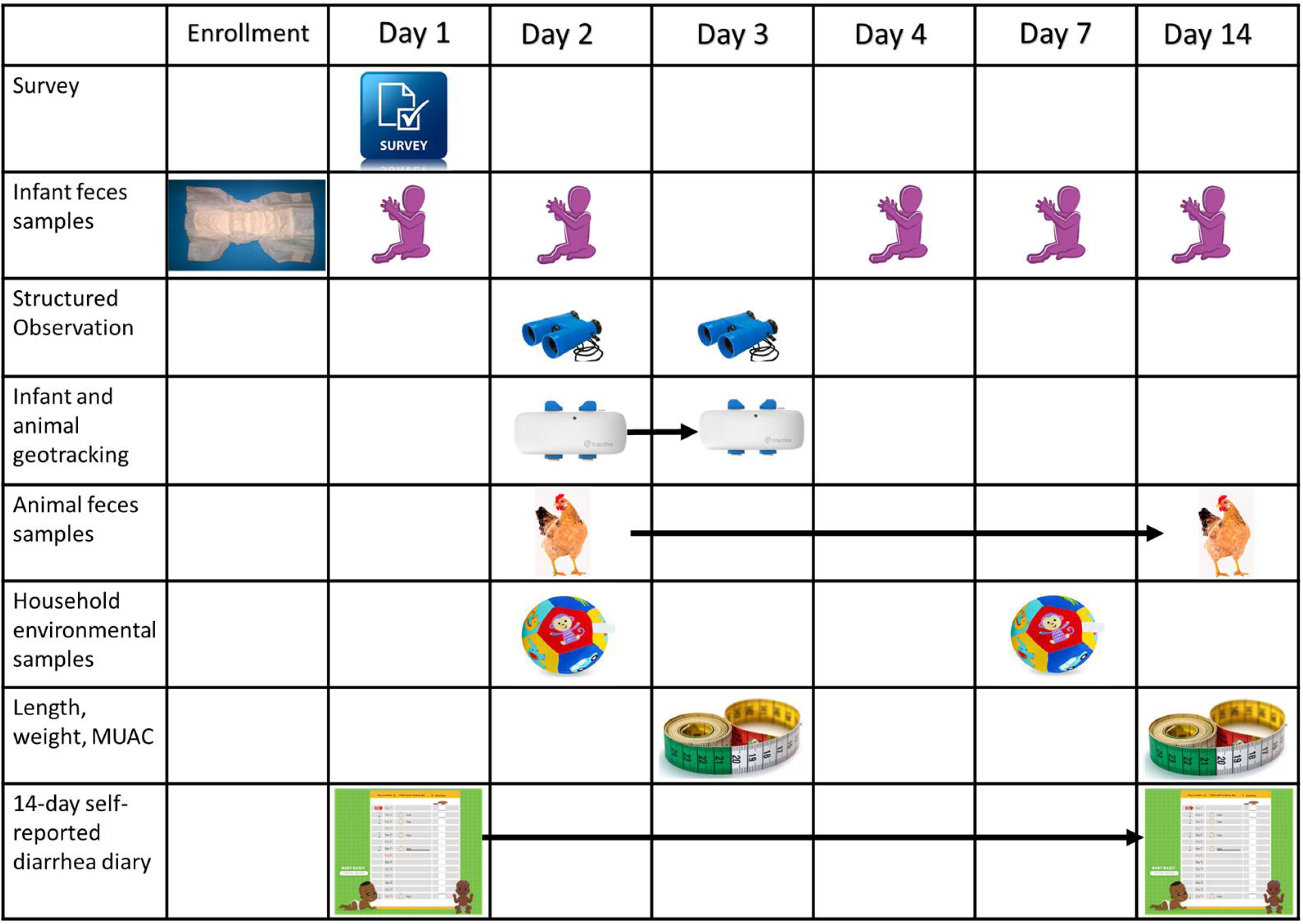
Study design and timeline for PATHOME household data and biological specimen collection. Symbols matching data collection activity placed under days when activities are implemented. Survey = self-reported data on household conditions. Diaper = provision of diapers for feces collection. Infant = days of diaper/feces collection. Chicken = animal feces collected. Binoculars = structured observation days. Geotracker = 24-hour time window where spatial movement is monitored. Toy ball = days of environmental sample collection. Infant on length board = anthropometric data. Calendar = window of time for prospective self-reporting of infant diarrhea symptoms. Photo credit for tape measure image: ShareAlike 3.0 Unported (CC BY-SA 3.0). Image of child licensed through Vectorstock Standard License. Other images taken by authors or are available through a Creative Commons license.

#### 6.1. Household Data Collection

Over the 14-day enrollment window households participate in a series of data collection activities as shown in **Error! Reference source not found**.3. On Day 1, our field staff collect the baseline diaper with infant feces and conduct a survey on socioeconomic conditions, behaviors, and health in the household. Caregivers are also given a 14-day self-reported diarrhea symptom calendar that spans enrollment to study completion to record diarrheal symptoms for the infant. On Day 2, field staff return for the first day of exposure assessment activities involving a five-hour period of structured observation, the placement of a geotracking device on infants and their domestic animals (if available), and collection of domestic animal feces and household environmental samples. The caregiver is reminded to continue using the diapers to collect infant feces and to record symptoms, if any, on the calendar. The geotracker is left on infants and animals for a 24-hour period spanning the morning of Day 2 to mid-afternoon on Day 3. On Day 3, field staff repeat 5-hour structured observation, collect a Day 3 feces specimen, document anthropometric measurements of the participating infant, and retrieve the geotracking devices just before departure. Field staff return to the household on Days 5, 7, and 14 to collect three more feces specimens (5 feces samples total). On day 7, the second anthropometric measurements are also documented. A second household soil/swab is also collected on this day. On Day 14, the enumerators return to the households to collect the diarrhea calendar, the last infant stool sample and provide caregivers the compensation gift.

##### 6.1.1. Household Survey

On Day 1, caregivers of infants are asked to respond to a 45-minute survey that documents demographics of persons living in the household, number and type of caregivers, wealth assets, household and compound infrastructure including access to drinking water sources and latrines, ownership and management of domestic animals, breastfeeding and feeding practices, method of handling infant and animal feces, handwashing practices, 7-day history of diarrhea and other symptoms in the infant and other household members and related health care utilization or self-administered treatments (e.g. antibiotics). Developmental indicators were chosen for their ability to serve as culturally relevant and discriminatory measures of upward socioeconomic mobility in Kenya.

##### 6.1.2. Prospective 14-day diarrhea calendar

The caregiver is given a 14-day pictorial diary with images of a sick and healthy infant and asked to mark any days in the next two weeks when the infant experiences 3 or more loose water or bloody feces in a 24-hour period. Enumerators and CHVs review the calendar on scheduled visits and prompt the caregiver to complete the record. On Day 14, the calendar is replaced with a gift of a 12-month calendar.

##### 6.1.3. Infant feces Collection

Five feces samples are collected per subject [Days 1, 3, 5, 7, and 14, yielding a total of 1,240 infant feces samples (248 infants * five feces samples)]. If staff return as scheduled but no feces is available, they are allowed to return anytime in the next 24 hours (e.g., Day 4 for Day 3) to fulfill the scheduled feces sample plan. Infant feces is collected by providing the caregiver a pack of disposable diapers sized by the infant’s weight, and asking her to use them immediately and for the duration of the 14-day study, providing any containing infant feces to the study team [55]. Diapers are used to collect infant feces to protect the feces from contamination by soil (e.g., scooped from ground) or feces of other children (e.g., potties in use by multiple children). If the infant defecates, the caregiver places it in a provided sterile, Ziploc bag and stores it in a safe, shaded place. The field team collects any diapers with feces during visits in a cooler with ice packs and transfers them to a supervisor for transport to a central lab for analysis (APHRC lab in Nairobi and Maseno University lab in Kisumu).

##### 6.1.4. Infant and Domestic Animal Spatial Geotracking

Data on the spatial-temporal movement of infants and domestic animals between household and public areas, and frequency and duration of co-localization that could lead to infant-animal interactions is measured by placing GPS mobile data loggers on infants and animals for a 24-hour period [63, 64]. The detailed development and validation of the geotracking protocol for infants and animals will be summarized in a separate manuscript. In brief, on Day 2 enumerators place a small (28 × 11-inch, 1.23 ounce) GPS Tractive Pet Tracker (Tractive Co., Austria) linked to an online real-time logging account on the infant. Enumerators work jointly with caregivers to identify an unobtrusive place for attaching the geotracker to infants, typically a snug child-sized wrist or ankle band or in a pocket of the infant’s clothing. Caregivers are assured that the devices do not record audio or visual information [65]. They are asked to move the location of the geotracker as necessary to ensure child comfort, minimize concerns of theft or visibility, changes in clothes, or other concerns, while keeping the tracker within arm’s reach of the infant [66, 67]. This tracker is placed prior to structured observation to allow enumerators to monitor infant discomfort or reactivity over the five-hour observation window and assist with placement fit strategies if necessary. This also provides parallel observation and satellite-based location data for examining the validity of methods for recording human behavior. The geotrackers remain placed for 24 hours, although if the caregiver choses to remove it from the infant for sleeping, she is asked to place the device next to, but out of reach of the infant and then replace the device when the infant wakes up.

An additional one or two geotrackers are attached to harnesses (chicken/duck, cat) and collars (dogs, goats, sheep, cow) of domestic animals belonging to the enrolled household. Enumerators monitor the animal for species-specific signs of distress for 15 minutes, and then turn the device on, which begins real-time upload of date, time, and longitude and latitude data to the web. If the animal does not adjust to the harness, then the harness is removed, and another animal, if available, is selected for observation. If the household owns multiple animals, staff prioritize placing geotrackers on two different species that most commonly come into contact with the infant over two animals of the same species or animals kept apart from infants and household areas. Geotrackers are set to transmit location data to satellites in real-time at five-minute intervals. The geotrackers are recovered after 24 hours on Day 3 for cleaning, recharging and reuse. Data is extracted from the Tractive server via an api and merged by geotracker ID with other household data for cleaning and summarization of movement and location of the wearer. A selected number of caregivers will be selected from the study sites for in depth interviews about their experiences with the geo tracking exercise. These caregivers will be randomly selected from households where tracking has been done for both animals and infants. The tool will contain questions about their experiences with the tracking, experiences of the infants and animals, placement of the tracker, their fears/concerns, and recommendations for improvement.

##### 6.1.5. Structured Observation of Infant and Caregiver Behaviors

After placing the geotrackers, enumerators begin 5 hours of structured observation of the enrolled child and their caregiver using a pre-piloted tool developed by our team using the Livetrak (developed by Stanford University) app for tablet data collection [42, 45, 65]. Tools document where the child is taken (public and household settings), what the infant is doing (e.g., sleeping, crawling, standing), and the rate of the child touching and mouthing soil, objects, food, drinking water, surface water, animals or their feces, and hands or other body parts of other children and caregivers. Tools also document the frequency of mitigating behaviors performed for infants, such as cleaning after defecation and infant feces disposal, cleaning face or hands, and hand washing [42]. In cases of multiple caregivers for children, the caregiver who is interacting with the child at any given time is recorded to capture differences in behavior by caregiver [54]. Data will be summarized by frequency of different observations as well as the sequence of behaviors.

##### 6.1.6. Domestic animal feces collection

Up to two specimens of animal feces representing different animal species are collected in each household, equating to 496 animal feces samples (248 households * 2 animal feces per household). Animal feces could include feces observed in animal enclosures if a household-owned animals or feces on the ground that may have been from personal or neighbor-owned animals. Enumerators will attempt to sample feces on Day 2, at the same time as observations, geotracking, and environmental sample collection. However, if no feces are observed on Day 2, enumerators can inspect the household on subsequent days when they return to collect infant feces and then collect samples. If no animal feces are observed over the 14-day time period, then the household will be considered uncontaminated by domestic animal feces. Feces specimens are collected into sterile WhirlPak bags, from the center of the pile to avoid contamination by soil, surface water, or other animal feces. These samples are stored in a cooler on ice packs and transported, as stated above, to the central lab for analysis.

##### 6.1.7. Household Environmental Samples

In each participating household, we collect two soil samples (Day 2 and 7), one caregiver hand rinse, one toy rinse, and in Kisumu only, one drinking water sample. Soil samples are about a 30 gram composite of soil from different locations in the household or a swab of a 30X30 cm squared floor space [68]. Toy rinses aim to measure the amount of microbial contamination that can be transferred onto household fomites in 24 hours. This is measured by providing the caregiver an alcohol sterilized infant toy, such as teething ring or rattle, on Day 1 for the infant to play with for 24 hours [69]. This infant toy is retrieved 24 hours later on Day 2 into a sterile barcode labeled bag for transport to the lab for microbial testing. An identical replacement toy is given to the infant for permanent keeping. At the end of behavioral observation on Day 2, one caregiver hand rinse is collected by submerging each hand into a barcode-labeled sterile one-liter WhirlPak bag with Phosphate Buffered Saline (PBS) and massaging all surfaces of the hand for 30 second before switching to the next hand. In total, we expect to collect 496 household soils, 248 toy rinses, and 248 hand rinses across all neighborhoods and cities, plus up to 124 drinking water samples (when water is consumed directly or indirectly via food preparation) in Kisumu. All samples are stored on ice packs and transported to the APHRC (Nairobi) or Maseno University (Kisumu) lab for processing and microbial analysis on the day of collection.

##### 6.1.8. Infant birth status and anthropometry data

Preterm birth status is recorded by extracting gestational week of age from the infant’s national birth registration booklet if possible, using caregiver self-report if the booklet is not available. Infant weight at the point of enrollment is measured using a digital scale. Infant length is measured using length boards per standard WHO guidelines [70]. These measurements are transformed into Z scores and categorized as wasted and stunted based upon whether their Z score is <2 standard deviations below international standard for weight and length for an infant of the same gender and weeks of age. If the infant’s booklet contains a record on infant weight and length at birth, this information is also recorded and used to calculate the absolute difference in weight and length from birth to enrollment age per week for comparison of average growth rates by city, neighborhood status, and other factors.

#### 6.2. Neighborhood Data Collection

##### 6.2.1. Environmental hygiene observations and soil and water collection

One neighborhood location is identified every four weeks in each study neighborhood using feedback from field staff as to locations where infants enrolled in the past four-week cycle were observed outside of their private households. If no infants left their homes during structured observation in the last four weeks, we consult the geotracker data to identify locations where the tracker placed upon the infant was found outside of household property. Repeating this process each month (12 months) in two neighborhoods in two cities results in 48 neighborhood areas where enrolled infants may have been exposed to neighborhood soil and water across a seasonal year. The designated locations are characterized by recording the types of urban infrastructure present, the presence of observed human waste disposal (diapers, flying toilets, individual piles, emptied buckets), and the presence of animal feces. Infrastructure includes small to large wastewater drains, paved roads, residences, electricity lines, community parks, businesses, and waste dumps. During neighborhood environmental observations, eight soil samples and up to four surface water samples are collected at each site to assess for pathogen contamination. The eight soil samples and four water samples are collected approximately 2m apart as previously described [39].

##### 6.2.2. Seasonality and weather data

Climate events like precipitation can cause small to large scale flooding that carries pathogens across the environment and contributes to their persistance in the environment. Middle class neighborhoods may have better infrastructure for preventing and managing run-off, so land-based precipitation estimates are likely inaccurate for measuring differences in urban flood exposure. A supplemental grant to the parent grant is supporting the development of strategies to generate neighborhood-specific satellite data on cumulative precipitation and flooding from multiple satellite-based hydrometerological online platforms over the 12 month time of data collection in each city for assessment of seasonal contributions to pathogen detection patterns.

### 7. Sample Processing and Microbial Assays

The overall sample processing strategy is shown below and in Figure 4 with detailed methods in Supplemental Materials.

**Figure 4.**
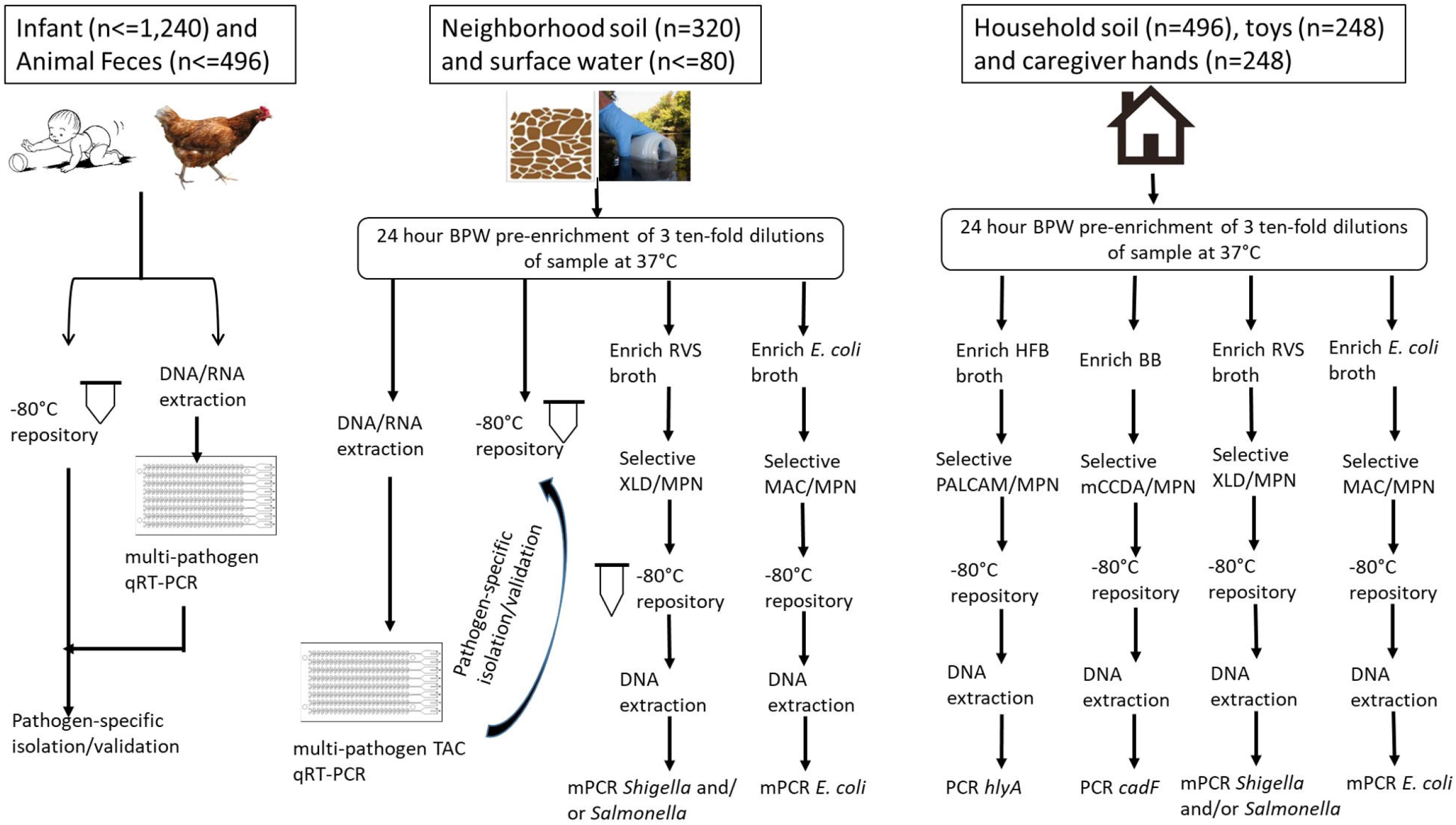
Flow diagram of the strategy for recovery, detection, quantification, and confirmation of enteric pathogens in human and animal feces and environmental fomites. Buffered Peptone water (BPW); Rappaport Vassiliadis Soya Peptone Broth (RVS); Half Fraser Broth (HFB); Bolton Broth (BB); modified Xylose Lysine Deoxycholate (mXLD); quantitative reverse transcription polymerase chain reaction (qRT-PCR); multiplex PCR (mPCR).

#### 11.1. Feces Processing

In a biosafety cabinet, a 300 mg portion of each feces specimen is transferred from the diaper into DNA/RNA Shield™ Collection Tubes containing ceramic beads and Shield cell lysis buffer (Zymo Research Corp, California, USA), spiked with a 10^5^ concentration of an extrinsic control for monitoring extraction efficiency, and extracted using the ZymoBIOMICS Quick DNA/RNA kit (Zymo Research Corp, California, USA). An extrinsic negative control tube of Shield buffer is processed twice a week to monitor for potential laboratory contamination. Approximately half of the Zymo tube is filled with feces and vortexed with an equal amount of Cary Blair preservation media (HiMedia, India) and 80% glycerol and stored in a biorepository in an ultralow freezer at the central laboratories. Some frozen feces samples are cultured for specific bacterial pathogens using the protocols described below to isolate individual bacteria for further validation of molecular assays.

#### 11.2 Quantitative molecular detection of enteric pathogens

DNA and RNA from infant and animal feces is analyzed as previously described using microfluidic TaqMan Array cards containing primers and probes for Norovirus GI and GII, adenovirus 40/41, *Sapovirus, Enterovirus*, Rotavirus, *Salmonella enterica, Shigella/EIEC backbone, Shigella/EIEC plasmid, Campylobacter jejuni/coli, Listeria monocytogenes, Clostridium difficile, Helicobacter pylori*, enterotoxigenic *Escherichia coli* (ETEC) LT/ST, enteropathogenic *E. coli* (EPEC), enteroaggregative *E. coli* (EAEC), and Shiga-like toxin-producing *E. coli* (STEC) stx1/stx2, *Enterocytozoon bieneasi, Giardia, Cryptosporidium spp*., *Entamoeba histolytica*, and the MS2 extrinsic control [3, 41, 71]. The TaqMan assays are run on a QuantiStudio 12K Flex Real-Time PCR System (ThermoFisher, Chicago, IL).

Prior to using the TaqMan Array cards, the assays are validated using ten-fold serial dilutions (10^0^ to 10^5^ genes per sample) of positive controls. Bacterial positive controls are prepared by culture of bacterial strains maintained at the University of Iowa. Other positive controls were prepared using qBlocks gene fragments (IDT, Coralville, IA, USA) with pathogen-specific sequence inserts targeted by the primer and probes. One molecular water PCR control for every box of 25 cards and two negative extraction controls per 100 samples is included to verify no contamination is introduced during sample preparation. Two field process negative controls per week are collected on days when samples are placed into Zymo tubes. These results are examined to verify MS2 extrinsic controls are positive but other genes are negative. Any feces samples processed on the same day as controls with false positive amplification are repeated by reisolating and extracting feces from the biorepository at the central labs. Inhibition is assessed by comparing cycle threshold (Ct) of MS2 to known concentration spiked into feces prior to extraction, plus calculating 260:280 and 230:260 ratios on a Nanodrop. The recovery efficiency is calculated by dividing the quantity of MS2 by the quantity of MS2 spiked into the sample prior to concentration.

#### 11.3. Selective Detection and Quantification of Environmental Bacteria

Presence and concentration of *Salmonella, Shigella, Campylobacter, Listeria, and E. coli* spp. in household soils, hand rinses, toy rinses, drinking water (Kisumu only), and neighborhood soil and water is measured using methods adapted from ISO and FDA food microbiology guidelines and relevant literature, and a Most Probable Number method that involves primary and secondary pre-enrichment and selective culture of serially diluted sample volumes.

First, three different volumes of soil, hand rinse, toy rinse, and water are measured into Buffered peptone water pre-enrichment broth (BPW, Himedia, India). Except for public surface water samples, all samples are subjected to 1:10, 1:100, and 1: 1000 dilutions of the original sample by measuring 25, 2.5 and 0.25 (g/ml) of samples into 225, 250 and 250 ml BPW, respectively. For water samples, assuming high likely chance of harboring diverse and high bacterial load, first a 10-fold dilution (1ml sample: 9ml BPW ratio) of public water surface sample is made from which subsequent serial dilutions (1:100 and 1:1000) are performed by serially transferring 1ml from the first dilution into tubes containing 9ml of BPW to yield 1:100, and 1:1000 dilutions. All the three dilutions are incubated at 37°C for 24h to recover sub-lethally injured bacteria.

Second, a 100 μL aliquot from each of the three BPW primary enrichment culture is transferred into 10 mL of selective secondary enrichment media using Rappaport Vassiliadis Soya Peptone Broth (RVS, Himedia, India) for *Salmonella and Shigella spp*., *E*.*coli* broth (Oxoid, UK) for *E. coli* spp., Bolton Broth (BB, Oxoid, UK) for *Campylobacter* spp., and Half Fraser Broth (HFB, Oxoid, UK) for *Listeria monocytogenes*. These are incubated at a growth temperature recommended for each pathogen (e.g. *Salmonella spp*. at 41.5°C for 24h).

Third, a loopful of each ten-fold sample dilution in secondary enrichment broth culture is streaked on selective agar and grown overnight to isolate specific strains, including steaking RVS on modified XLD (mXLD, Oxoid, UK) for *Salmonella and Shigella* spp., *E. coli* broth on Eosin methylene blue or MacConkey agar (Oxoid, UK) for *E. coli*, BB on mCCDA (Oxoid, UK) for *Campylobacter* spp., and FB on ALOA (Oxoid, UK) for *Listeria monocytogenes*. These are incubated at 37°C for 24h. Incubation of *Campylobacter* spp. occurs in anaerobic chambers with gas packs (Mitsubishi gas Chemical America Inc., Japan) except for the primary enrichment.

Fourth, up to ten representative colonies of each distinct phenotype are collected from each plate, sub-cultured into tryptic soya broth (Himedia, India) and incubated at 37°C for 24h and then preserved as glycerol stocks at 1:2 ratio for future re-isolation and analysis. For DNA extraction, a loopful of aliquot from the glycerol stocks are sub-cultured into Tryptic Soy agar/selective agar and then five to ten representative colonies are transferred into 100 μL molecular grade water, boiled for 10 minutes, centrifuged to precipitate cell debris, and the supernatant transferred to a new sterile labeled Eppendorf tube. DNA samples from presumptive positive bacteria are stored at −20° C until pathogen status can be confirmed by qRT-PCR. Identity of each type of presumptive enteric bacteria phenotype is verified by polymerase chain reaction assays for gene indicators of pathogen-specific identity of isolated colonies. Concentration of presumptive bacterial species is determined by examining which sample cultures across the three volumes of environmental sample tested for each bacterial protocols contained in presumptive positive colonies using Most Probable Number (MPN) method as described in Appendix 2 of the Food and Drug Administration Bacteriology Analytical Manual [72].

#### 11.4. qRT-PCR Verification of environmental bacteria species

Pathogen status of presumptive *Salmonella, Shigella, Campylobacter, Listeria*, and *E. coli* spp. isolated from household soil, hands, toys, and neighborhood soil and water is tested by qRT-PCR of DNA isolated from bacterial colonies growing on tryptic soya agar/selective agar. The PCR is conducted by distributing 18 μL of mix (10 μL of master mix, 1 μL of TaqMan assay containing specific primers and probes and 7 μL of nucleic acid-free water) first and then, 2ul of DNA template of the bacterial isolate to each well in a 0.2 ml 96-well PCR plate, alongside PCR negative controls. The qRT-PCR primers and probes and the PCR cycling conditions are identical to those used on TAC to ensure comparability of detection information.

### 14. Biorepository

A biorepository generated through this study is preserving all human and animal feces specimens, bacterial isolates from selective culture of environmental samples, and the non-selective buffered peptone broth primary enrichment cultures of those environmental samples for validation of results as well as for future research. Primary stool specimens and bacterial isolates from the environment are stored in an ultralow freezer (−76°C) at the stated central labs and DNA/RNA/cDNA isolated from all specimens is stored in an ultralow freezer (−76°C) at the University of Iowa.

### 14. Analysis

Primary data analysis has been described on clinicaltrials.gov, and will involve characterizing and modeling differences in socioeconomic, infrastructure, behavioral, spatial, environmental, zoonotic, and human data per our study design: by city and low versus middle-income neighborhood type, and by age group. The form of analysis and presentation will be determined by data type and distribution (e.g., principal component analysis of wealth, diversity and network mapping of pathogen data). Estimates of pathogen exposure dose and diversity in pathogen type per exposure pathway will be performed as previously described [43]. Many statistical methods and agent-based models are being developed newly through this study and will be publicly disseminated via publications, open data sharing resources, and on our own websites.

### 15. Community engagement and Translational Science

Community and stakeholder engagement by the research team occurs prior to study onset in each city to gather feedback from community leaders, health workers, policy makers, and other academic and non-governmental stakeholders in each city. Suggestions for improvement or concerns about risks to participants are used to improve study protocols. Feedback also contributes to a sense of ownership of the study by local policy makers and practitioners, ensuring that results will be received and potentially implemented and sustained after the study to address local health challenges. Community and stakeholder engagement is repeated at the end of the study to share initial learning from the studies, and a final end-of-study meeting at the end of the statistical modeling and virtual lab design stage to disseminate final recommendations. To ensure research is quickly and effectively translated into policy, the research team is conducting a landscaping review of Government of Kenya policies across sectors, including urban housing and development, health, and agricultural to identify opportunities for improving existing policies, implementing policies that address gaps, or linking policies between sectors for improved planning and monitoring.

### What this study will contribute

This study will be able to identify the most significant pathways of enteric pathogen transmission in urban low-to-middle income settings in Kenya. Based on the new learning we will be able recommended new or modified strategies for combining interventions to expedite enteric disease control for settings like urban Kenya. Unique elements of the study as described in our Rationale include a focus on <u>patho-microbial community (PATHOME)</u> dynamics for examining pathogen transmission patterns across exposure pathways and rigorous microbiological tools for characterizing the pathome. We recruit <u>middle class households</u> with high basic standards of development <u>and</u> use a <u>14-day repeat sampling cohort design with</u> integrated household and neighborhood microbial exposure assessment. Our combined behavioral observation and newly developed geotracking methods provide rigorous new evidence about <u>spatial dimensions of exposure</u>, in terms of where infants experience environmental exposures and frequency and duration of localized interaction with domestic animals. Our Planetary Health-One Health integrated approach includes documentation of urban socio-economic development as well as domestic animal health and their potential to serve as zoonotic vectors through direct or indirect interaction with humans. The data generated through this cohort study will be used to construct new statistical and agent-based models to identify spatial-temporal dynamics in enteric pathogen transmission in LMICs and the role of societal development in preventing transmission. An interactive virtual laboratory will be developed to predict how different improvements in household and neighborhood development alone and in combination could influence endemic enteric disease burden in Kenya or other urban settings. The overarching goal of this virtual laboratory is to generate targeted recommendations for improving policy and practice, especially in terms of preventive options, and provide a system for refining those interventions over time in the context of ongoing societal changes. We will publicly share the large datasets we collect, thereby providing an invaluable asset to other modeling groups working on reducing incidence rates of enteric pathogen infections in children in LMICs.

## ETHICS AND DISSEMINATION

This study conducts research on both human and animal health outcomes. The protocols for human subjects’ research for the PATHOME study were approved by Institutional Review Boards at the University of Iowa (ID -202004606), AMREF Health Africa (ID -ESRC P887/2020), and a permit obtained from the Kenyan National Commission for Science Technology and Innovation (ID# P/21/8441). The study is also registered on Clinicaltrials.gov (Identifier: NCT05322655). Protocols for research on canines, felines, avians, and ruminants were approved by the UI Animal Care and Use Committee (ID 0042302) in accordance with the Guide for the Care and Use of Laboratory Animals, NIH Publication No. 85-23 (2010). Details on human and animal research ethical considerations and protocols are reported in Supplemental Material. The entire research team took refresher training on ethical principles on protecting human research participants.

## Data Availability

All data produced in the present study are available upon reasonable request to the authors

https://pathomelab.github.io/

## AUTHOR’S CONTRIBUTIONS

KKB and DKS conceived of the study and secured funding. KKB drafted the epidemiological study design, with input from DKS, SS, AZ, and BM. SS and PJB designed field protocols and survey, behavioral, diarrhea calendar, and anthropometry tools, with support from AZ, AO, and KKB. KKB and FDG designed field sampling and laboratory protocols, with input from BO, JA, CSA, AGK, and CO. BO, JA, CA, and CO implemented culture-based laboratory protocols, and FDG and AK implemented molecular and genomic protocols. PJB, AMO, and KKB designed geotracking protocols. PJB, SS, SG, and DKS oversaw data management and quality control. AZ, SS, and CO facilitated human subjects’ and animal research approvals and stakeholder engagement in Kenya. KKB oversaw human subjects and animal research protocol approvals in Iowa. BM conducted policy landscape analysis. KKB and FDG oversaw enteric pathogen analysis approach. DKS oversaw the statistical and agent-based analysis of the epidemiological data. KKB wrote the manuscript, with input from all authors.

